## Supplementary material for "Wound Complications Following Surgery to the Lymph Nodes: A protocol for a systematic review and meta-analysis": Search Strategy

### **Supplementary Table 2. Search strategy for ‘**Wound Complications Following Surgery to the Lymph Nodes: A protocol for a systematic review and meta-analysis’

Databases searched: EMBASE; Medline

| Procedure terms  Connected with an OR | Boolean Operator | Outcome terms  Connected with an OR | LIMITS |
| --- | --- | --- | --- |
| exp "SENTINEL LYMPH NODE BIOPSY"/  OR exp "LYMPH NODE EXCISION"/  OR "lymph* gland* biops*"  OR "lymph* node* biops*"  OR "lymph* tissue* biops*"  OR "lymph* gland* dissection*"  OR "lymph* node* dissection*"  OR "lymph* tissue* dissection*"  OR "lymph* gland* excision*"  OR "lymph* node* excision*"  OR "lymph* tissue* excision*"  OR "lymph* gland* extirpation*"  OR "lymph* node* extirpation*"  OR "lymph* tissue* extirpation*"  OR "lymph* gland* resection*"  OR "lymph* node* resection*"  OR "lymph* tissue* resection*"  OR Lymphadenectom*  OR Lymphoadenectom* | AND | exp "SURGICAL WOUND DEHISCENCE"/  OR exp SEROMA/  OR exp HEMATOMA/  OR exp "SURGICAL WOUND INFECTION"/  OR exp "POSTOPERATIVE COMPLICATIONS"/  OR "postoperative wound infection*"  OR "surgical infection*"  OR "surgical site infection*"  OR "surgical wound infection*"  OR " wound dehiscence"  OR "wound disruption*"  OR "wound rupture*"  OR "wound separation*"  OR "postoperative complication*"  OR "postsurg* complication*"  OR "surgical complication*"  OR Hematoma*  OR Hematoderma*  OR Haematoma*  OR Haematoderma*  OR seroma* | DT 1991-2021  Languages English  Humans |

Databases searched: CENTRAL

**#1** (Lymph*):ti,ab,kw AND (biopsy):ti,ab,kw AND (complication*):ti,ab,kw (Word variations have been searched) **#2** (Lymph*):ti,ab,kw AND (biopsy) AND (complication*):ti,ab,kw AND ("postoperative complication"):ti,ab,kw (Word variations have been searched)
